# A syringe-based digital algometer with a USB interface: a low-cost alternative to commercially available devices

**DOI:** 10.1101/2025.03.11.25323786

**Authors:** Stepan Frankevich, Aryeh Simmonds, Izhak Michaelevski, Daniel Yakubovich

## Abstract

Quantitative pain assessment is important for effective pain management. Pain pressure threshold (PPT) and Pain Tolerance (PT) measured through pressure algometry offer valuable tools for quantitative evaluation of nociceptive stimuli. Commercially available pressure algometers are often expensive, lack modularity and cannot integrate well into multi-component monitoring systems. Although low-cost algometers are available, most require complex calibration and lack a digital interface, limiting real-time data acquisition and integration with electronic health record systems.

In the current study, we describe a durable and accurate pressure algometer built on a syringe, an Arduino microprocessor and an analog piezoelectric pressure sensor. The PPT values obtained with our device align well with data obtained with commercially available digital and mechanical algometers. In addition, our device can be easily connected to computer via a USB, allowing for convenient data storage and analysis.

Our results demonstrate the accuracy and reliability of a novel algometry device constructed from readily available materials and requires minimal engineering and programming skills. The device operates without addition calibration and can seemlessly integrate with a computer using open-source software.

**Highlights:**

- We present a digital algometer comprised of a standard plastic syringe, an analog pressure sensor, and an Arduino UNO microcontroller.
- The device provides applied force measurement with high precision, compatible with commercially available algometers.
- The device is user-friendly, requiring no special skills or software to use, and transfers data directly to a computer. Additionally, it may be synchronized with other biosensors for multichannel signal acquisition.

## Introduction

According to epidemiological data, as much as 30% of the population regularly experience pain [1,2]. Exposure to acute pain dramatically increases risk of developing chronic pain, disrupts recovery after trauma and limits function of affected structures [3]. Painful experiences combine nociceptive signaling with additional emotional and cognitive components, making accurate assessment of pain difficult [4]. To address this, several methods for objective assessment of pain have been developed.

Algometry is one of the most widely implemented methods of pain assessment. The basic principle of algometry is application of a steadily increasing nociceptive stimulus until the sensation becomes painful (Pain Threshold), or until the pain becomes too distressing (Pain Tolerance). Many modes of stimuli may be utilized, including high and low temperatures and application of pressure [5].

Pressure algometers are commonly used for pain assessment in clinical settings and in research. These tools have demonstrated excellent reliability and accuracy [6]. Existing designs are typically based on calibrated springs and digital pressure sensors to calculate force applied to the patient. However, the high cost of commercially available algometers can limit their use. To address this issue, several inexpensive designs have been developed. A low-cost model based on a plastic syringe has been developed for use in an emergency response unit setting [7]. Another solution has been proposed in several studies – utilization of a force gauge / hand-held dynamometer as an algometer [8,9]. These devices share their working principle with algometers and demonstrate high validity and reliability.

In this study, we propose a digital, low-cost algometer based on a plastic syringe and a high precision pressure sensor. This device can provide high-resolution records of applied force, which can be simultaneously transferred to computer for further processing.

## Materials and Methods

Our device operates on a piston principle similar to a previously described tool [7]. In the current study, we used plastic syringes of various volumes combined with a T-type luer-lock valve in order to create an air-tight cylinder. The modified thumb rest of the syringe is utilized as a pressure probe. The overall design of the algometer is presented in **Fig. 1**.

**Figure 1.**
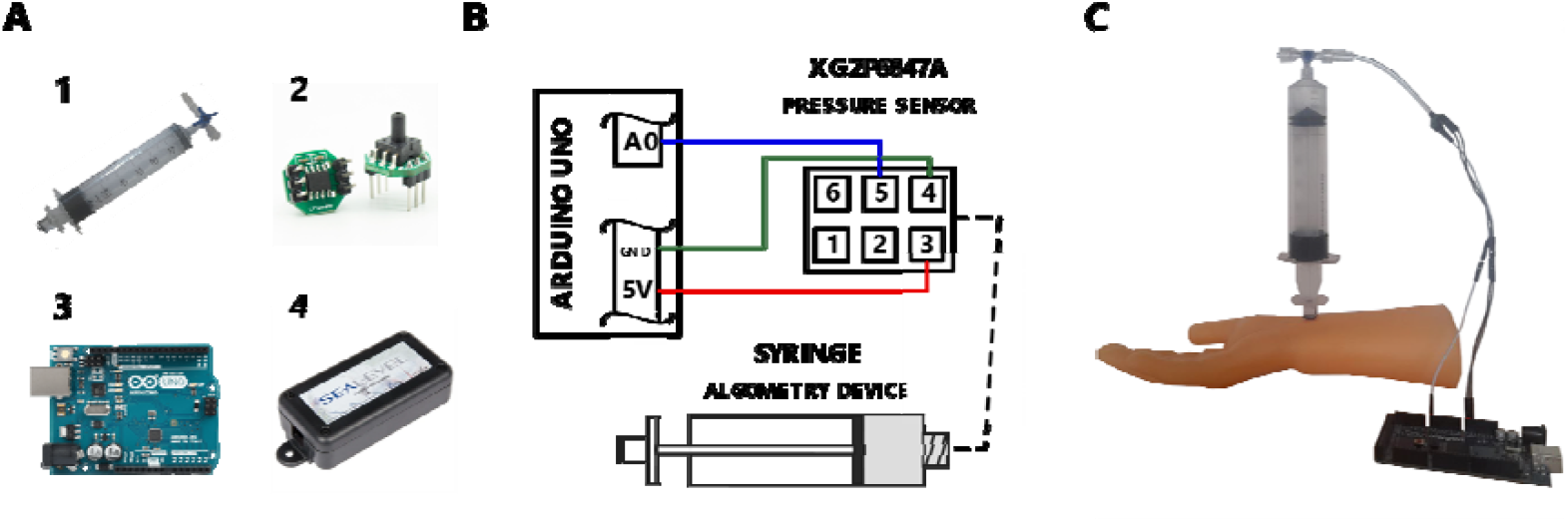
Design of the syringe-based digital algometer. **A**. Components of the syringe-based algometer. **1:** A plastic luer-lock syringe (Plastipak Luer-lock Syringe, Becton, Dickinson and Company (BD), United States) with a T-type luer-lock and a 1cm^2^ pressure probe. **2:** An XGZP6847A Analog Pressure Sensor. **3:** An Arduino UNO microprocessor. **4:** A SEALEVEL SealSO USB Isolator connected to the PC. **B**. A graphic guide for assembly of the syringe algometer. The dash line represents a plastic tube. **C**. The syringe-based algometer used on a model limb.

All further described modifications were performed for every syringe size. The syringe thumb rest has been flattened using abrasive paper. A 1 cm^3^ acrylic cube has been affixed to the thumb rest with epoxy glue, creating a pressure probe. The side of the probe in contact with the skin has been covered with a 1 cm^2^ sheet of rubber to prevent slipping. A luer-lock compatible T-type valve has been connected to the tip of the syringe. In this way, the syringe has been converted into a sealed chamber with volume and internal air pressure affected by plunger position. An 85 mm long, 3 mm wide plastic tube with 1 mm-thick wall has been connected to the syringe through the T-type valve. This tube served as a connector between the syringe and the pressure sensor.

In order to access concurrent validity of pressure measurements conducted with our device, we utilized the NUL-210 pressure sensor (SES Education, Israel) instead of XGAZP6847A piezoelectric pressure sensor (CFSensor, China).

For validation of force measurements, we implemented the NUL-225 force plate sensor (SES Education, Israel). We utilized a square wooden frame to maintain our device in fixed position. Using a manually operated lifting table, the Nul-225 pressure plate was gradually elevated, compressing the device and increasing the internal air pressure. For each syringe size, a set of equidistant volumes was predetermined. During each test, the internal pressure was gradually increased until the relevant volume was reached and maintained on that level until the end of the test. Each test lasted for 5 minutes **(see Fig.1S)**.

Nul-210 and Nul-225 sensors, which have built-in microcontroller, were directly connected to the computer via USB. Data from these sensors was recorded and saved in the comma separated value (CSV) format using the NeuLog Windows Application (SES Education, Israel). The XGAZP6847A piezoelectric pressure sensor (CFSensor, China) was connected to the Arduino microcontroller (Arduino, Italy). Data recording and transfer from Arduino microcontroller to the computer was performed using a script written in Aruino IDE (see Supplementary Materials). Time-pressure data series were acquired using the Putty Serial Console (an open-source tool) format for further analysis. Throughout the experiment, data output was visually monitored through the serial console. Specifications of all sensors used in this work are summarized in Supplementary Table 1. To assess concurrent validity of our device compared with commercially available algometers, we modified two existing algometers by attaching pressure tips identical to that used in our device (an acrylic cube with a 1 cm^2^ surface). The modified commercial algometers were the 60 Pound BASELINE Dolorimeter (Fabrication Enterprises, USA, algometer similar to Fischer [8]) and the AMF-500 Digital Force gauge (ALIYIQI, China). Pain Pressure Threshold (PPT) measurements were obtained from twelve healthy (7 female, 5 males), ages of 24 and 52, using our device and two modified algometers. Individuals reporting active pain, either acute or chronic, were not included. All tests were conducted in a calm environment, with only the participant and the researcher present. Each device was applied at adjacent points within the test area, located at the midpoint of the frontal forearm surface (Fig. 1C), three trials, with the 3-minute interval between tests. The pressure was gradually increased until the sensation of pressure shifted to minimal pain. Upon the participant’s verbal report of pain, the device was promptly removed, and the syringe’s plunger was manually extended to the maximum volume position. This procedure was repeated for the BASELINE and the AMF-500 algometers, with force readings recorded manually at the onset of pain.

All data are presented as mean ± standard error of mean. Concurrent validity of pressure and force measurements was estimated using linear regression calculated in SigmaPlot 11 for Windows (Grafiti LLC, USA). To estimate data bias, we used Bland-Altman plots [9].

All experiments involving human subjects were approved by the Ariel University Medical School Ethical Committee (approval number AU-MED-DY-20231219).

## Results

As a first step, we compared the results of pressure measurements obtained from the XGAZP6847A sensor and the NUL-210 pressure sensor. **Fig. 2** shows data collected using a 60 ml syringe, where the plunger was moved to a specific syringe volumes, and the corresponding pressure was recorded with both sensors. Additional data from the comparison of pressure sensors for syringes of other volumes (3, 5, 10 and 20 ml) are provided in Supplementary Materials, Figure 2s.

**Figure 2.**
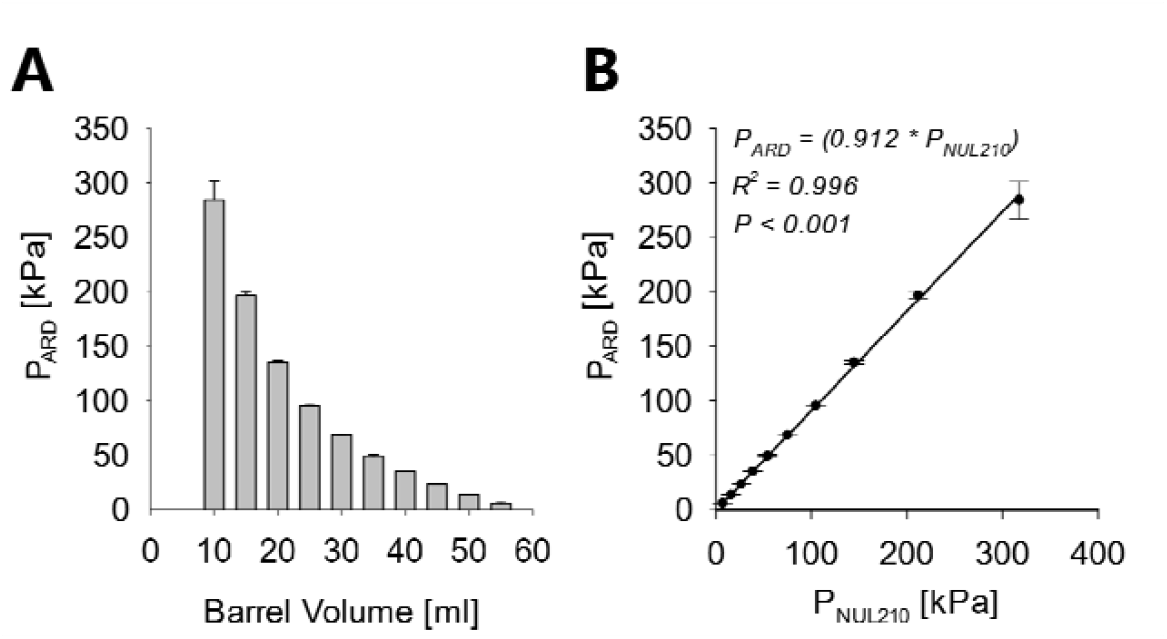
Pressure measurement validation. **A**. Pressure–volume relationship estimated for 60 ml syringe. Each bar represents averaged results from 5 separate measurements at each specific volume. Data obtained with XGAZP6847A pressure sensor. **B**. Averaged pressure measurements taken at each specific volume with the XGAZP6847A pressure sensor (P_ARD_) and the NUL-210 pressure sensor (P_NUL210_). Solid line represents linear regression. R^2^ and P-value stated on the plot (Spearman correlation analysis).

In order to assess the possible bias of pressure measurements, we utilized Bland-Altman analysis for each volume **(Fig. 3)**.

**Figure 3.**
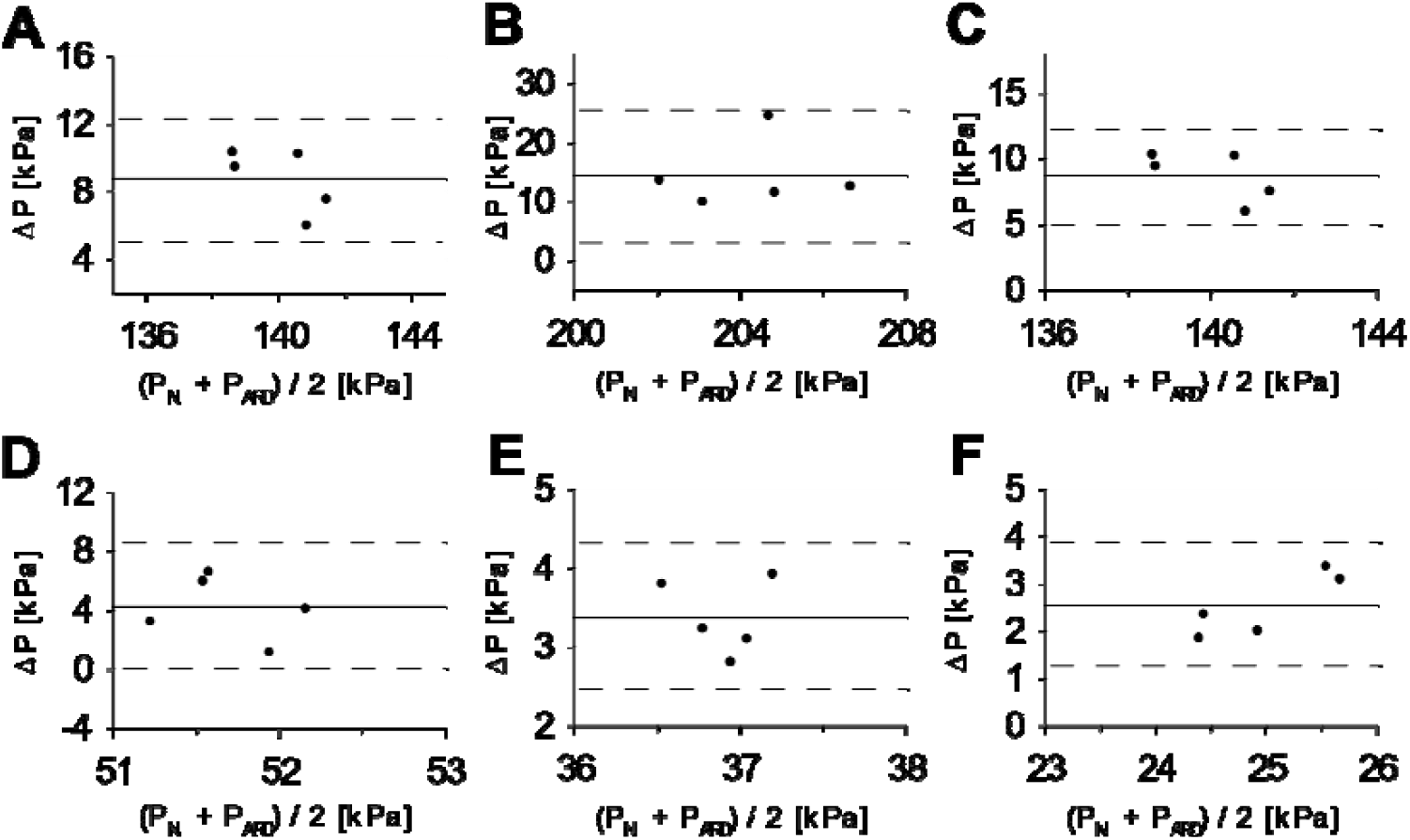
Comparison of internal air pressure recorded using the NUL-210 and XGZP6847A pressure sensors in the syringe-based algometer (60ml). **A**-**F**. Bland-Altman plots comparing measurements of internal air pressure made with NUL-210 (P_N_) and XGZP6847A (P_ARD_) pressure sensors at specific volumes. Each data point represents a separate test. **A:** 15 ml; **B:** 20 ml; **C:** 25 ml; **D:** 35 ml; **E:** 40ml; **F:** 45 ml. Δ*P = P*_*NUL210*_ *- P*_*ARD*_

We estimated the generated force for each pressure measurement based on Boyle’s Law [10] **(Fig.4 A)**. The probe area was calculated as the area of circle, with the diameter measured utilizing caliper (probes areas for different syringes are summarized in Table 2s). For comparison, we conducted force measurements using the NUL-225 force plate sensor, recording the generated force for each syringe volume where a pressure measurement was taken. The data are shown in **Fig.4 B-H**.

Subsequently, we compared the pressure threshold data obtained with our device to measurements taken with two commercial algometers. Data collected from a group of human volunteers is shown in **Fig. 5**.

## Discussion

In this study, we present a low-cost algometer constructed from a plastic syringe, a pressure sensor and an Arduino microcontroller. The performance of this device was extensively evaluated utilizing the Nul-210 pressure sensor and the Nul-225 force sensor. In both cases, correlation coefficients obtained from linear regression analysis were close to 1 (Pearson correlation test), as demonstrated in Fig 2B and Fig. 4B, indicating good concurrent validity with previously tested devices. Moreover, our device demonstrated high precision in internal pressure measurements, with a standard error-to-signal ratio of 0.1-3.4% (Fig.2A). Bland-Altman analysis of both pressure and force measurements showed no bias (Fig.3 and Fig.4). Based on these findings, we suggest that our device exhibits acceptable concurrent validity and precision compared to existing devices.

**Figure 4.**
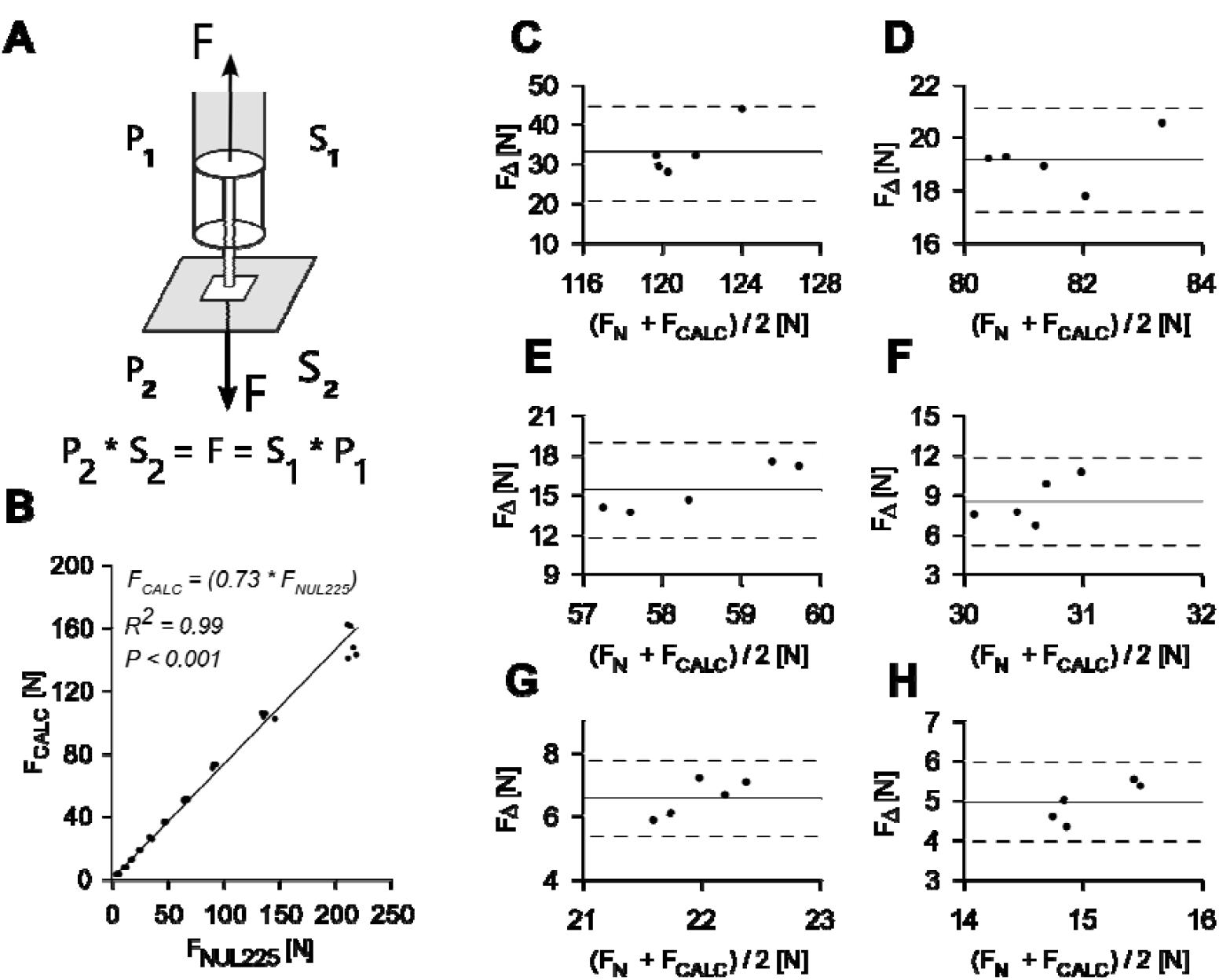
Comparison of directly measured force (NUL-225 Force Plate) and calculated force 60ml syringe. **A**. Schematic representation of force calculation from internal air pressure. The force generated by application of the probe to the skin can be calculated as either product of the pressure measured inside the syringe (P_1_ – parameter reported by our device) and surface area of the plunger (S_1_, which was measured for each syringe, see Table 2 Supplementary material). The value P_1_*S_1_ is supposed to be equal to the force generated by plunger tip on the surface of the skin (which is actually a product of P_2_ (pressure on the skin) and S_2_ (the area of the pressure probe)). **B**. Directly measured force (F_NUL225_) and force calculated from internal air pressure (F_CALC_). Solid line represents linear regressions in a 60ml syringe. **C-H**. Bland-Altman plots comparing directly measured force (NUL-225 Force Plate) and force calculated from internal air pressure (XGZP6847A pressure sensor) at specific volumes. **C:** 15 ml; **D:** 20 ml; **E:** 25 ml; **F:** 35 ml; **G:** 40ml; **H:** 45 ml. _Δ_*F = F*_*NUL225*_ *-F*_*CALC*_

Parenthetically, as shown in Fig.4B, while the correlation coefficient for the relationship between calculated and measured force is close to 1, the proportion coefficient is less than 1. This discrepancy may be due to the fact that the rubber syringe plunger is not a perfect flat circle, but rather a conic shape, resulting in a surface area larger than that calculated using the formula for a circle. Furthermore, direct comparison of pain threshold measurements utilizing our device with those obtained from two commercially available algometers demonstrated good agreement when tested on human volunteers (Fig.5A, 5C).

**Figure 5.**
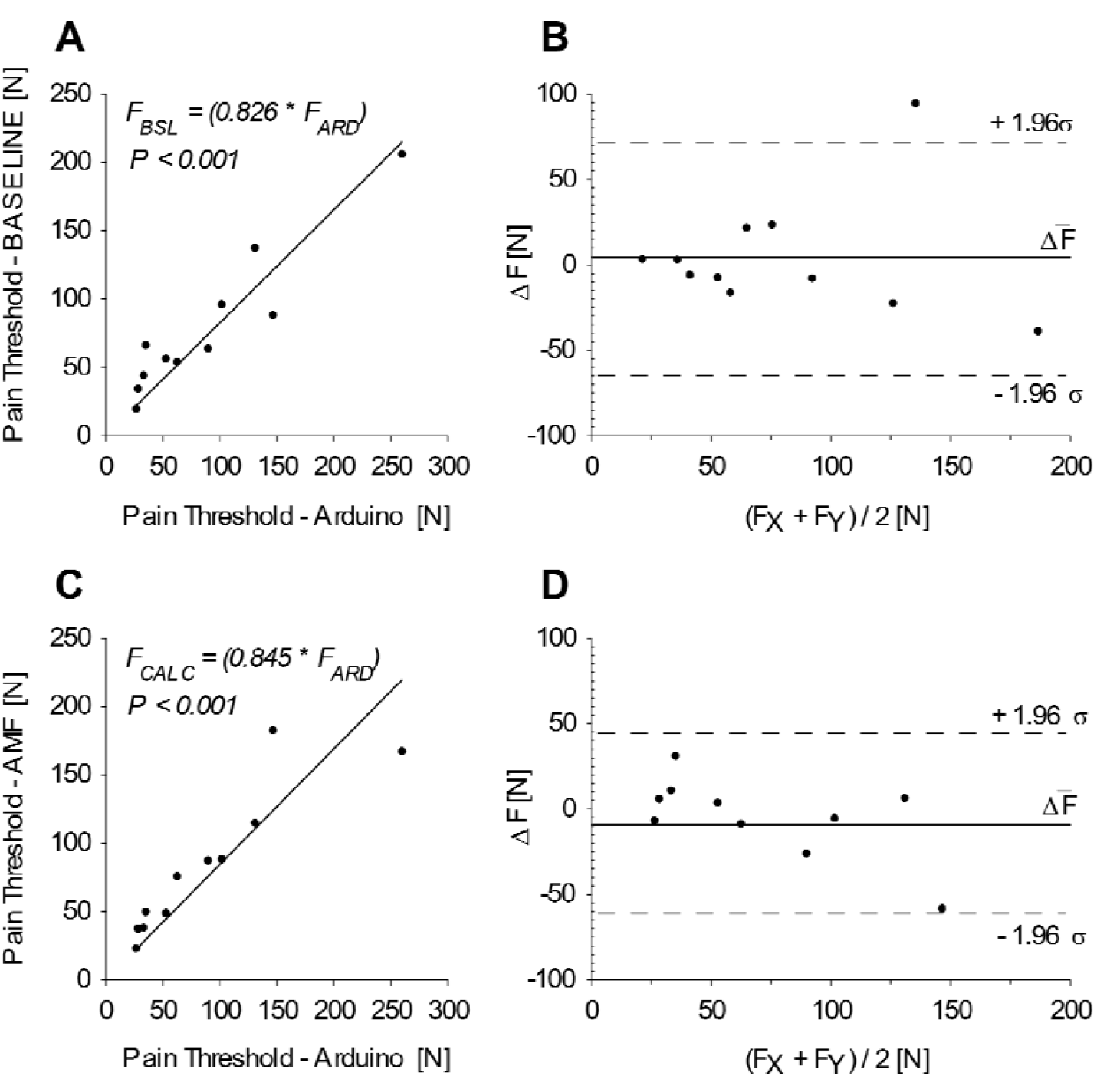
Comparison of the syringe-based algometer (60ml) with commercially availiable algometry devices. A Pain threshold measured with the syringe-based algometer and the BASELINE mechanical algometer. **B**. Bland-Altman plot comparing the syringe-based algometer and the BASELINE algometer. **C**. Pain threshold measured with the syringe-based algometer and the AMF digital algometer. **D**. Bland-Altman plot comparing the syringe-based algometer and the AMF digital algometer. Solid lines represent linear regressions.

As previously discussed, several digital algometers are currently available on the market, including the Algomed (Medok Instruments, Israel), Commander Echo (MTM, Canada), Wagner FPIX (Wagener Instruments, USA) among others. However, these devices come with significant costs, typically ranging from 900 USD to several thousand USD, making them financially prohibitive for newer or smaller laboratories [11]. In contrast, the total cost of our device, including the sensor, syringe, connectors and tubing - remains below $150. As of now, all components are readily available for international purchase, offering a more affordable alternative.

The development of syringe-based algometry devices has been explored in previous studies, such as for short-term use in emergency response units [7] or for examination of coccydinia [11]. The primary advantages of our device lie in its integration of an analog pressure sensor and Arduino microprocessor. This combination addresses the calibration challenges that have been a limitation of earlier syringe-based algometers [7]. It also enables high frequency data sampling (up to 15 kHz) and supports multi-channel data acquisition. As demonstrated in the attached script, the programming requirements are minimal (see Supplementary material). Moreover, in the future the multi-channel acquisition capability of our device could be expanded to connect several biometric sensors (*e*.*g*. electrodermal activity, ECG, EMG) to the same microcontroller.

In summary, the current study describes an affordable, low cost, and high precision algometry device designed for measuring Pain Pressure threshold and Pain tolerance. With future development and necessary certification, this device has the potential to be implemented as a medical device in the future.

## Data Availability

All data produced in the present work are contained in the manuscript

## Abbreviations

PPT: pain pressure threshold
VAS: Visual Analog Scale
NRS: Numeric Rating Scale
MPQ: McGill Pain Questionnaire
ADC: Analog-Digital Converter

## Supplementary Materials

**Table 1s.**
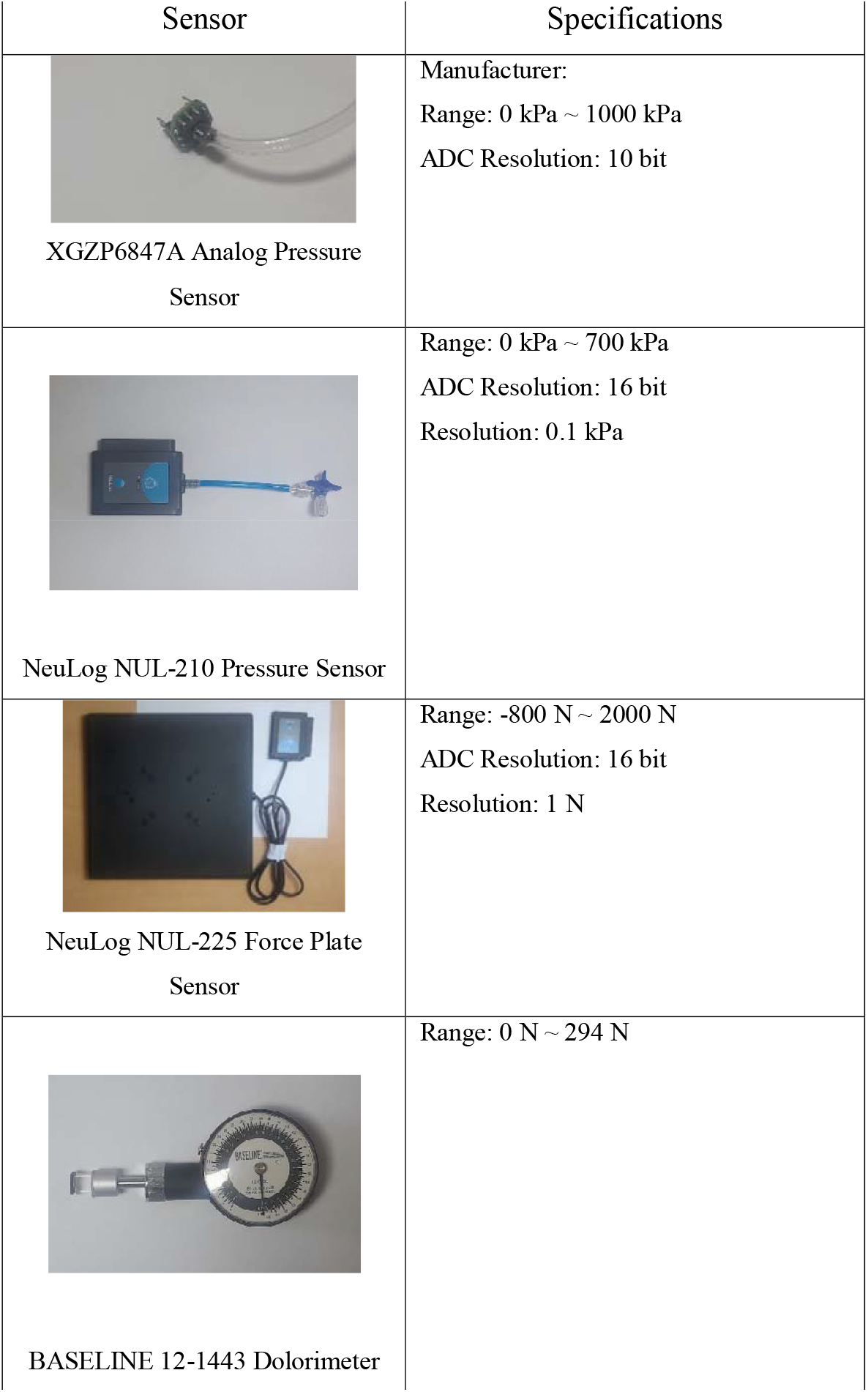
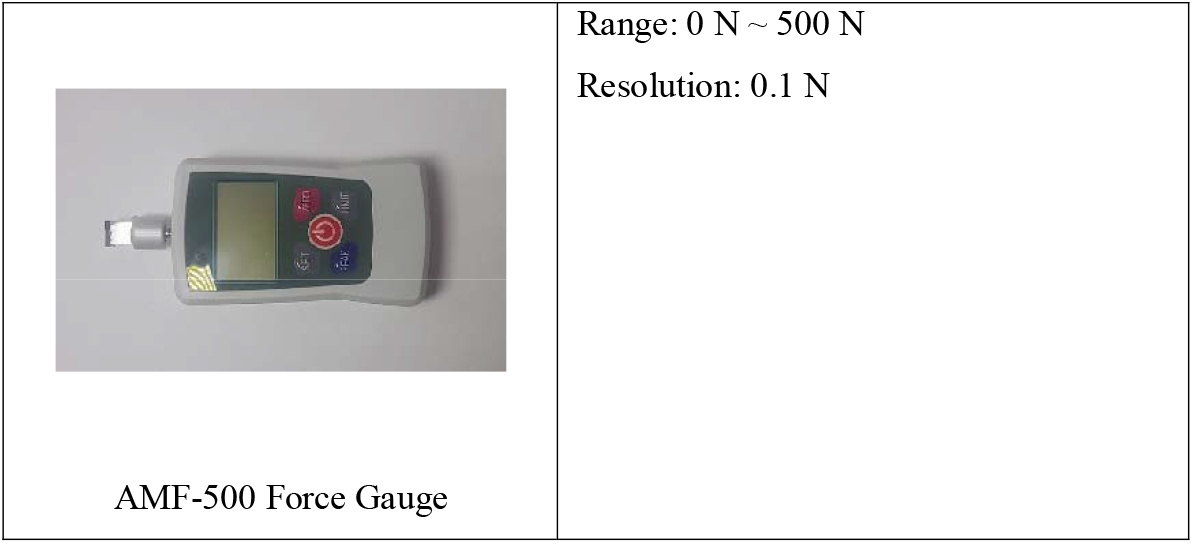
Technical specifications of sensors and algometry devices involved in the study.

|  |  |
| --- | --- |
| Sensor | Specifications |
| XGZP6847A Analog Pressure Sensor | Manufacturer:<br>Range: 0 kPa ~ 1000 kPa<br>ADC Resolution: 10 bit |
| NeuLog NUL-210 Pressure Sensor | Range: 0 kPa ~ 700 kPa<br>ADC Resolution: 16 bit<br>Resolution: 0.1 kPa |
| NeuLog NUL-225 Force Plate Sensor | Range: -800 N ~ 2000 N<br>ADC Resolution: 16 bit<br>Resolution: 1 N |
| BASELINE 12-1443 Dolorimeter | Range: 0 N ~ 294 N |
| AMF-500 Force Gauge | Range: 0 N ~ 500 N<br>Resolution: 0.1 N |

**Table 2s.** Surface area of rubber plunger lining (S_1_ on Fig.4A)

| Table 2s. Surface area of rubber plunger lining ($S_1$ on Fig.4A) | |
| --- | --- |
| Syringe total volume (ml) | Surface area of the circular base of the cone used in calculations [$m^2$] |
| 3 | $0.59 \cdot 10^{-4}$ |
| 5 | $1.14 \cdot 10^{-4}$ |
| 10 | $1.65 \cdot 10^{-4}$ |
| 20 | $2.87 \cdot 10^{-4}$ |
| 60 | $5.6 \cdot 10^{-4}$ |

**Figure 1s.**
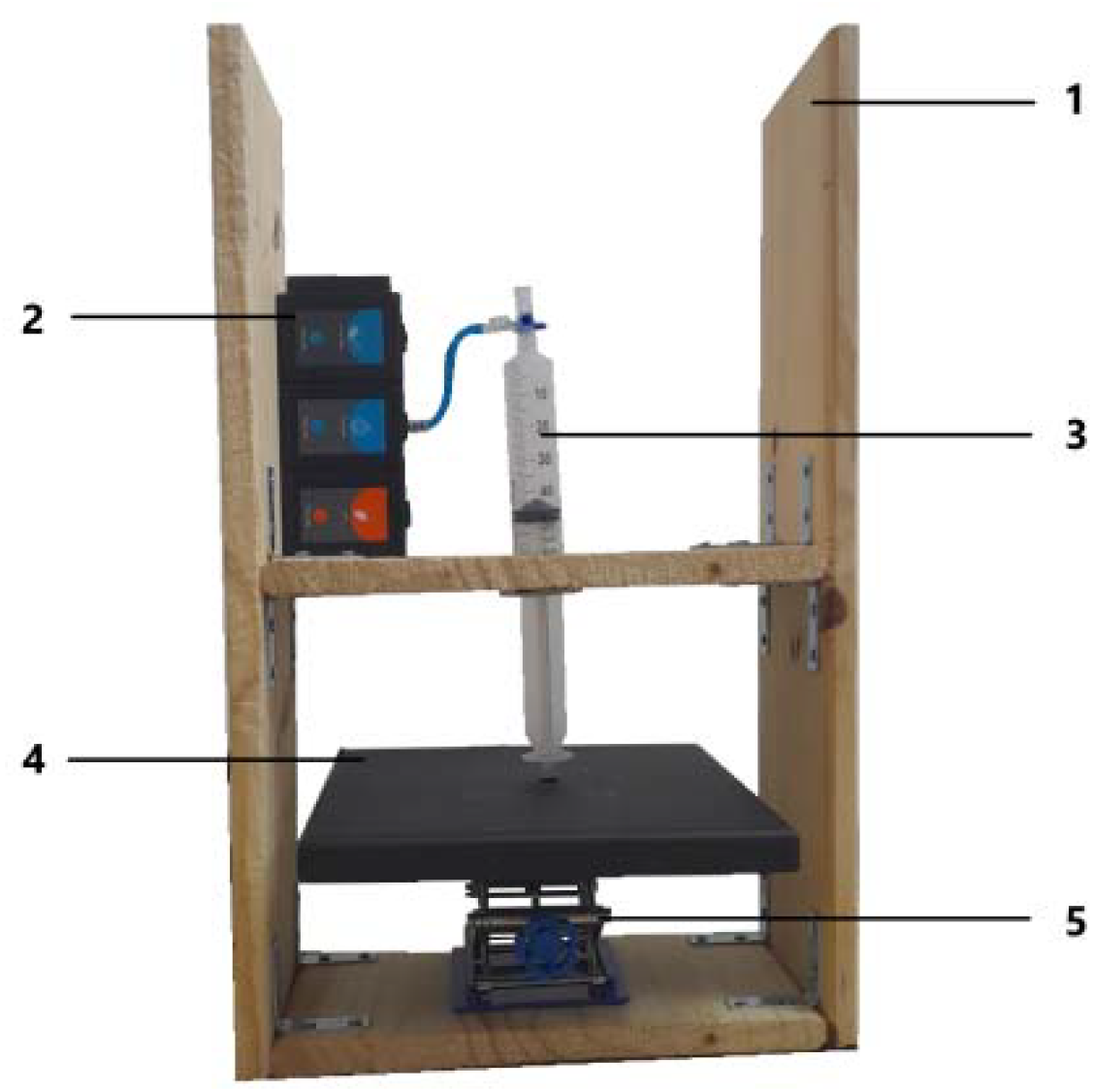
Validation of force measurements against internal air pressure. **1:** A wooden frame was constructed consisting of two vertical wooden panels connected with two perpendicular, shorter panels with metal corner braces and metal wood screws. A round opening was created in the center of the middle panel, wide enough to fit the syringe, leaving the flanges below that panel. **2:** The NUL-210 pressure sensor connected to the NUL-225 force gauge and the USB module. The pressure sensor connects to the syringe through an airtight luer-lock. **3:** The sealed syringe. A pressure probe was affixed to the thumb rest. The flanges help hold the syringe in place during the test. **4:** The sensor pad of the NUL-225 force plate. **5:** A manual lifting table. During the test, the lifting table is extended, pushing the force plate up and compressing the air inside the syringe. Resulting changes in the internal air pressure and force exerted onto the force plate are monitored and recorded.

**Figure 2s.**
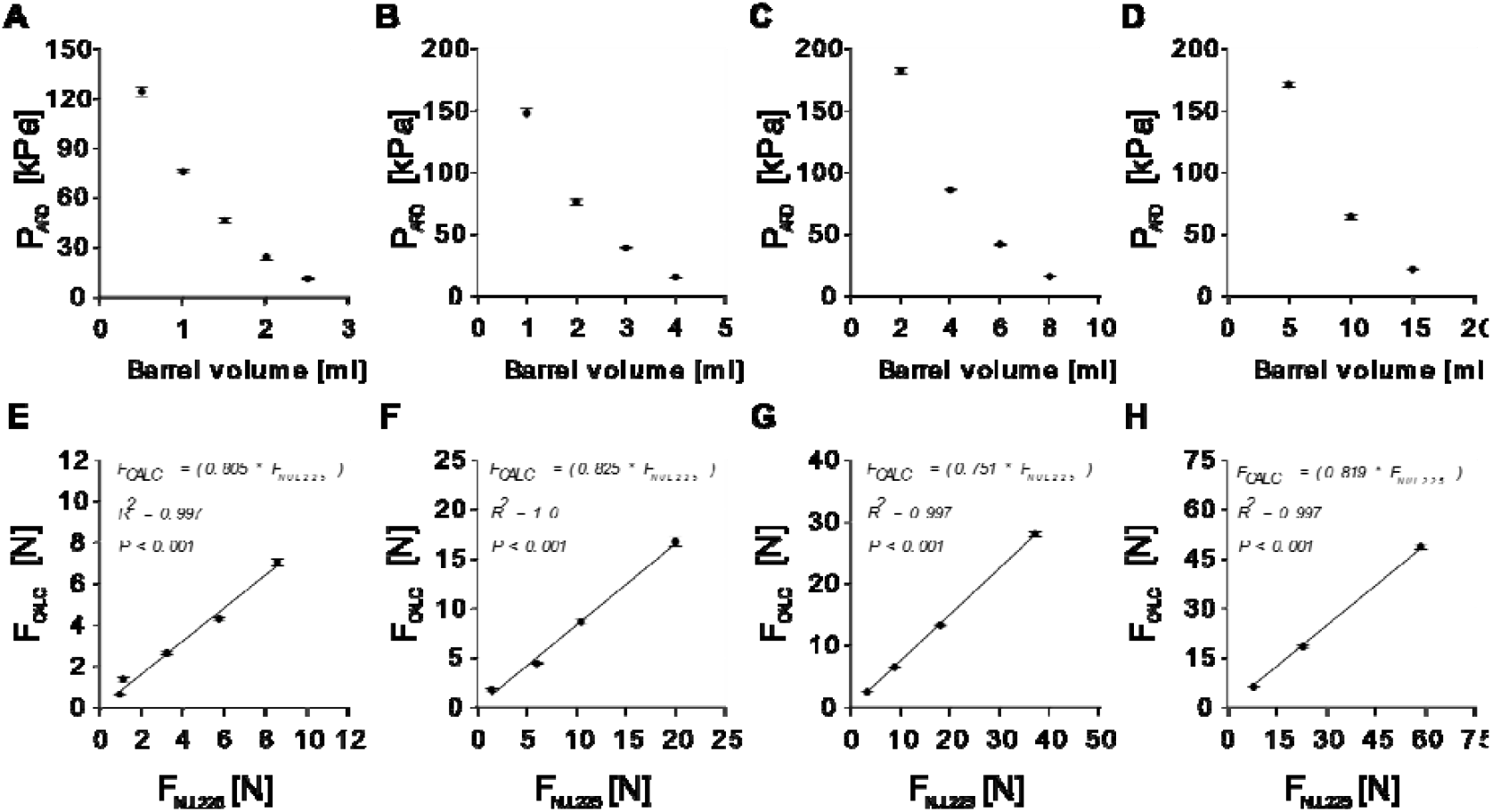
Precision of the syringe-based algometer (3ml, 5ml, 10ml, 20ml) in repeated tests. **A-D**. Averaged XGZP6847A pressure sensor measurements at specific volumes. **A:** 3 ml; **B:** 5 ml; **C:** 10 ml; **D:** 20 ml. **E-H**. Linear relationship between expected force values calculated from measurements of internal pressure and force applied to a horizontal surface of the NUL-225 force plate. **E:** 3 ml; **F:** 5 ml; **G:** 10 ml; **H:** 20 ml.

### Arduino IDE script for acquisition of data

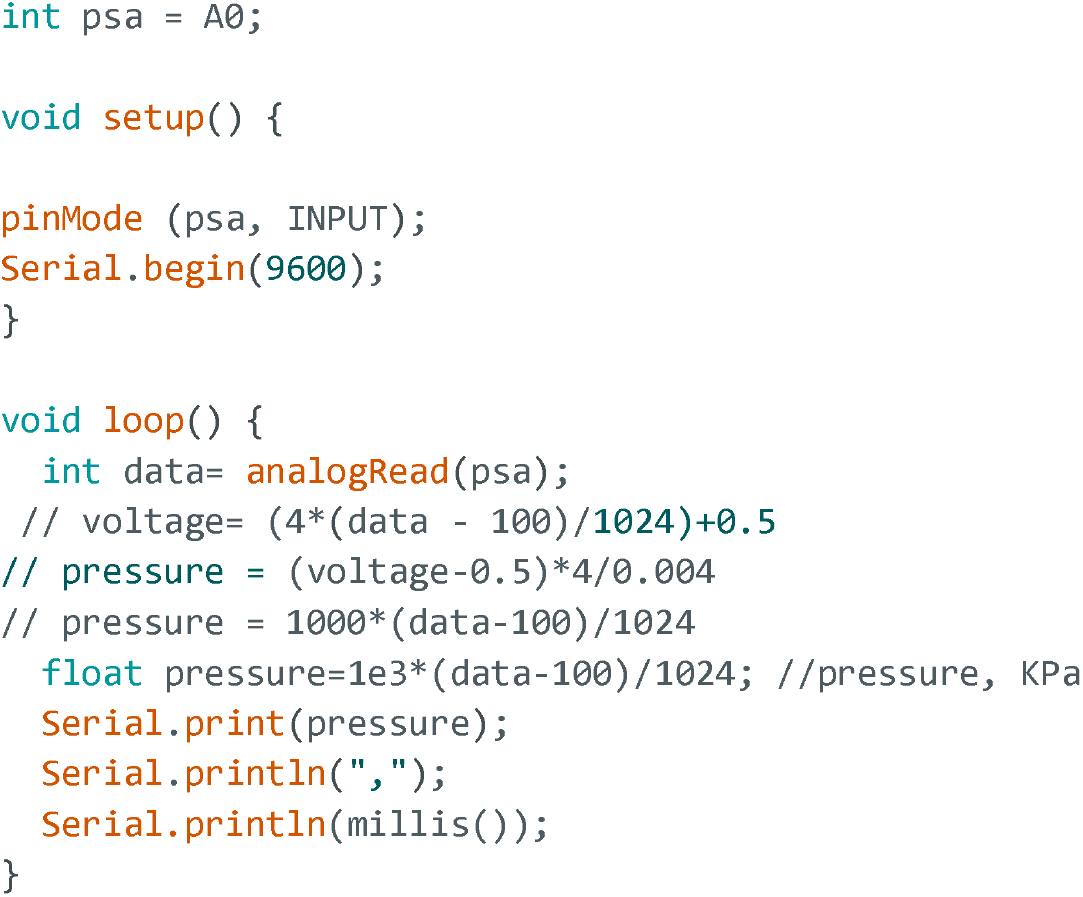

The Arduino script was designed to repeatedly calculate the internal air pressure (Pa) from the sensor 10-bit output. In order convert the measured value of pressure in KPa into applied force (N) we utilized Eq.1:

Eq.1 F = pressure _arduino·1e3·S

where F is calculated force, pressure_arduino is measured pressure and S is the surface area of the pressure probe (m^2^). The probe areas are summarized in Tab.2s.

